# Performance on verbal fluency tasks depends on the given category/letter: Preliminary data from a multivariable analysis

**DOI:** 10.1101/2021.12.30.21268567

**Authors:** Petar Gabrić, Mija Vandek

**Author notes:** <u>Corresponding author:</u> Department of Psychiatry and Psychotherapy, Philipps University of Marburg, Rudolf-Bultmann-Straße 8, 35039 Marburg, Germany.

## Abstract

Verbal fluency tasks are often used in neuropsychological research and may have predictive and diagnostic utility in psychiatry and neurology. However, researchers using verbal fluency have uncritically assumed that there are no category-or phoneme-specific effects on verbal fluency performance. We recruited 16 healthy young adult subjects and administered two semantic (animals, trees) and phonemic (K, M) fluency tasks. Because of the small sample size, results should be regarded as preliminary and exploratory. On the animal compared to the tree task, subjects produced significantly more legal words, had a significantly lower intrusion rate, significantly shorter first-response latencies and final silence periods, as well as significantly shorter between-cluster response latencies. These differences may be explained by differences in the category sizes, integrity of the categories’ borders, and efficiency of the functional connectivity between subcategories. On the K compared to the M task, subjects produced significantly more legal words and had significantly shorter between-cluster response times. Counterintuitively, a corpus analysis revealed there are more words starting with ⟨m⟩ compared to ⟨k⟩ in the experimental language. Our results potentially have important implications for research utilizing verbal fluency, including decreased reproducibility, questionable reliability of diagnostic and predictive tools based on verbal fluency, decreased knowledge accumulation, and increased number of publications with potentially misleading clinical interpretations.

## 1. INTRODUCTION

### 1.1. Verbal fluency in basic and clinical neuroscience

Verbal fluency (VF) is a common neuropsychological tool used for assessing cognitive and linguistic abilities in both healthy and clinical populations (Oberg & Ramírez, 2006; Pekkala, 2012; Stielow, 2017; Thiele et al., 2016; Turkstra, 2018). Most commonly, the semantic (SF) and phonemic fluency (PF) tasks are administered, in which the subject is to name as many words of the given semantic category (e.g., *animals*) or beginning with the given letter/phoneme (e.g., *F*), respectively, in limited time. Usually, only the productivity score (i.e., the raw number of legal words) is calculated.

The relative pervasiveness of SF and PF tasks in neuropsychological research can at least partly be attributed to their simplicity and brevity which increase the probability that the subject will understand the task and maintain the instructions in their working memory throughout the task. This makes VF particularly suitable for use in psychiatric and neurological populations where the presence of psychopathological phenomena might hinder understanding of the task and its completion (Holmlund et al., 2019). Furthermore, the task requires continuous behavioral responses from the subject, making it appropriate for studying brain metabolism during cognitive tasks via neuroimaging techniques (Costafreda et al., 2006; Gonzalez-Burgos et al., 2021; Kircher et al., 2011; Nagels et al., 2011; Scheuringer et al., 2020; Vonk et al., 2019; Wende et al., 2012). Both SF and PF have been used for studying functional brain metabolism in diverse psychiatric disorders, including depression, schizophrenia, bipolar disorder, etc. (Backes et al., 2014; Corrigendum, 2015; Erratum, 2021; Husain et al., 2020; Takizawa et al., 2014; Tassi et al., 2022; Yeung et al., 2021), as well as in studies where psychiatric symptoms are induced in healthy subjects, e.g., via infusion of subanesthetic ketamine (Nagels et al., 2011a, b). VF tasks are further used for establishing the presence of specific cognitive deficits in pathological populations (Stielow & Stenneken, 2017). Deficits in the productivity on SF and LF in relation to healthy subjects have been used as arguments for deficits in semantic memory, executive functioning, linguistic processing, etc. (Aita et al., 2019; Amunts et al., 2020, 2021; Kavé et al., 2020; Ledoux et al., 2014; Shao et al., 2014; Ventura et al., 2005; Whitesite et al., 2016) in pathological populations such as patients with mild cognitive impairment (Bauer et al., 2021), Alzheimer’s disease (Henry et al., 2004), Parkinson’s disease (Henry & Crawford, 2004c; Højlund et al., 2017), Huntington’s disease (Henry et al., 2005), traumatic brain injury (Cermak et al., 2021; Henry & Crawford, 2004a, b; Rosenkranz et al., 2020; Thiele et al., 2016), multiple sclerosis (Santangelo et al., 2019), epilepsy (Metternich et al., 2014), schizophrenia (Bokat & Goldberg, 2003; Henry & Crawford, 2005a; Rosenkranz et al., 2019; Tan et al. 2020, 2021; cf. Gabrić, 2021a, b), depression (Henry & Crawford, 2005b; Klumpp & Deldin, 2010), bipolar disorder (Raucher-Chéné et al., 2017), obsessive-compulsive disorder (Henry, 2006), autism (Spek et al., 2009), specific language impairment and other developmental disorders (Mengisidou, 2019), HIV infection (Iudicello et al., 2006), and many others.

Furthermore, there is an increasing number of studies suggesting that different behavioral and neural measurements of both SF and PF may diagnostically specific for different types of pathologies and/or discriminate between healthy individuals and patients of different pathologies, including cognitive impairment and dementia (Jacobs et al., 2020; Lam et al., 2006; Linz et al., 2017; McDonnell et al., 2019; Radanovic et al., 2007, 2009), schizophrenia (Becker et al., 2009; Costafreda et al., 2011; Takizawa et al., 2014), bipolar disorder (Costafreda et al., 2011), localization of cerebral lesions (Pendleton et al., 1982; cf. Turkstra et al., 2005; Wong et al., 2010), and others. VF may be used to discriminate between mild cognitive impairment and dementia (Bertola et al., 2014; Zhao et al., 2013), between vascular cognitive impairment without dementia and dementia (Zhao et al., 2013), and between idiopathic normal pressure hydrocephalus and dementia (Louta et al., 2017). Several studies have shown that VF tasks may be used as tools for early prediction of cognitive impairment and dementia (Clark et al., 2016; Cosentino et al., 2006; Gomez et al., 2006; Pakhomov et al., 2018; Sutin et al., 2019; Tröger et al., 2018; Vaughan et al., 2018).

VF tasks are also used within language lateralization assessments (Birn et al. 2010; Bishop et al. 2009; Knecht et al. 2000; Meyer et al. 2014; Uomini et al., 2013) and they are used as a measure of verbal intelligence (Crawford et al., 1992; Miller, 1984).

### 1.2. Category-specific and letter-specific effects on verbal fluency performance

Most researchers utilizing SF and PF uncritically assume that there are no category-specific or phoneme/letter-specific effects on the quantity and quality of performance on different SF and PF tasks. This is evidenced (1) from the fact that different studies draw the same inferences about cognitive deficits or other phenomena using different semantic categories and/or letters and (2) from the fact that studies administering multiple semantic categories and/or letters typically calculate only the aggregate score for the different SF and/or PF tasks. Despite this silent and uncritical assumption present in much of the literature, there are no data to support this assumption and, more problematically, there are clear indications in the linguistic and neuropsychological literature that differences in performance patterns on different semantic categories and letters should be expected. Given that different performance patterns might reflect different underlying cognitive processes (Vandek et al., 2018) and that VF tasks are often used for exploring cognitive processes in pathological populations, the silent assumption of SF and PF homogeneity might have important implications for research using SF and PF tasks and might explain some reproducibility issues in research using VF.

Neuroscientific research has clearly demonstrated that semantic memory is widely distributed over the cortical surface and the processing of different semantic categories may be associated with disproportionate levels of metabolic activity in different brain regions (Dreyer & Pulvermüller, 2018; Fahimi Hnazaee et al., 2018; Gabrić, 2019, 2021c; Gainotti, 2015; Huth et al., 2016; Liuzzi et al., 2020). Furthermore, both theoretical and experimental linguistics agree that different lexical-semantic features such as concreteness/abstractness and animacy/inanimacy underlie divergent cognitive processing mechanisms, while pathological populations may exhibit feature-specific deficits on lexical tasks (Carammaza & Shelton, 1998; de Almeida et al., 2021; De Letter et al., 2019; Seijdel et al., 2021; Sekulić Sović et al., 2019). Similarly, processing of different phonological categories is associated with different cognitive and neural mechanisms (Kronrod et al., 2016; Lago et al., 2015).

Despite this, the effect of the given category or phoneme/letter on performance on SF and PF has been alarmingly understudied. It is clear from studies reporting descriptive data of multiple SF tasks that there are stark discrepancies in the number of legal words produced by healthy subjects across different semantic categories. Typically, adult healthy subjects produce the most words on animal fluency, while productivity on other semantic categories may be reduced by more than twofold (Bosanac et al., 2017; Gabrić & Vandek, 2021a; Jebahi et al. 2020, 2021; Kim et al., 2015; Moreno-Martínez et al., 2017; Vicente et al., 2021). For example, Jebahi et al. (2020) reported that subjects produced 18 words on average on animal fluency but less than ten on the categories *accessories, tools, transportation means, furniture*, and *sports*. One study found that performance on categories *animals* and *vegetables* was associated with neither age nor education, unlike *fruits* (Farghaly et al., 2018). Moreover, pathological populations may show deficits of different degrees on different SF categories or they may show normative deficits on one but not the other category. One study on SF in Alzheimer’s disease has revealed that the extent of the deficit is dependent on the given category’s size (Diaz et al., 2004), while Moreno-Martínez et al. (2017) and Neves et al. (2020) found that although the animal fluency is by far the most commonly utilized SF task for research on dementia, other categories such as *supermarket items, fruits*, and *vegetables* are more sensitive in distinguishing healthy people from patients with dementia. Gabrić and Vandek (2021a) reported that inpatients with first-episode psychosis displayed deficient clustering on the categories *animals, vegetables*, and *musical instruments* but not on *trees* and *fruits*, despite the presence of a normative deficit in productivity across all categories, while Tagini et al. (2021) reported that patients with Parkinson’s disease showed semantically disorganized output on the category *fruits* but not *animals*.

Similarly, there is a long-standing division between “easy” and “difficult” letters in English within PF assessments (Borkowski et al. 1967), recently supported by Barry et al. (2008) who reported from a meta-analysis that performance on the CFL version (i.e., letters *C, F*, and *L*) of the Controlled Oral Word Association Test (COWAT) was significantly reduced and thus more difficult compared to the FAS version, as well as that healthy subjects exhibit greater variability on the FAS compared to the CFL version (cf. Ferrett et al., 2014).

The eventual existence of category- and phoneme/letter-specific effects on SF and PF performances may have important implications for the past and current research using VF, including decreased reproducibility, questionable reliability of diagnostic and predictive tools based on verbal fluency, decreased knowledge accumulation, and increased number of publications with potentially misleading clinical interpretations. Although there exist normative values for VF performance which are category-or letter-specific, they are not always used for interpretation in psychiatric and neurological research and in almost all of the cases, they relate to the raw number of legal words the subject produced, ignoring thus a number of other more specific aspects of VF performance.

The aim of the present study was to investigate whether there are differences in performance patterns within different SF and LF tasks. We compared performances in multiple variables between two SF and two LF tasks.

## 2. MATERIALS AND METHODS

### 2.1. Subjects

Sixteen healthy subjects were recruited for the study (6 males, mean age 22.250 ± 2.543 years, mean education 15.188 ± 1.759 years). All were native speakers of one of the Croatian dialects, students at the University of Zagreb, young adults, and right-handed as assessed by the Edinburgh Handedness Inventory (Oldfield, 1971). Subjects who had more than one native language or had a history of psychiatric or neurological disorders were not included in the study. Ethical approval was obtained by the Ethical Committee of the Faculty of Humanities and Social Sciences, University of Zagreb (dated 2018/10/03). All subjects signed an informed consent form.

### 2.2. Verbal fluency assessment

Subjects were to name as many words according to the given cue in 60 seconds. The categories *animals* and *trees* were used for SF, while the letters ⟨k⟩ and ⟨m⟩ (corresponding to phonemes /k/ and /m/ in Croatian dialects) were used for PF. An exploration of the Croatian web corpus hrWaC (Ljubešić & Erjavec, 2015) revealed that there were more lemmas starting with ⟨m⟩ (N = 436 719) compared to ⟨k⟩ (N = 412 961) (Goodness of fit: χ (1)^2^ = 664.300, p < .00001). PF was administered one to two weeks after SF to avoid SF-related recency effects on PF. Responses were audiotaped and transcribed using ELAN (Version 5.8) (ELAN, 2019; Wittenburg et al., 2006). Clusters were identified according to Troyer et al. (1997), with the cluster threshold set to 2 words (Abwender et al., 2001; Lehtinen et al., 2021; Troyer & Moscovitch, 2006). Two native speakers independently rated the outputs. Discrepancies were discussed until mutual agreement was achieved. Clustering and switching variables were calculated according to Gabrić and Vandek (2021a).

Dependent variables included: 1) legal words (raw), 2) perseveration rate (%), 3) intrusion rate (%), 4) first-response latency (ms), 5) final silence (ms), 6) cluster magnitude (words per cluster), 7) switching rate (%), 8) between-cluster response latencies (ms), and 9) within-cluster response latencies (ms).

### 2.4. Statistics

Statistical analyses were performed in JASP (Version 0.11.1.0) (JASP Team, 2020). Comparisons between the animal and tree variables, and K and M variables were performed using the paired-sample Wilcoxon signed-rank test. The intrusion rates on SF were compared with the one-sample Wilcoxon signed-rank test with the test value set at 1 ± 1, as no subjects produced intrusions on the animal task. The effect sizes are given by the matched rank biserial correlation. Correlations were explored using Spearman correlation coefficients.

## 3. RESULTS

### 3.1. Comparisons and correlations within semantic fluency tasks

Results from the comparisons are displayed in Table 1. On the animal compared to the tree task, subjects produced significantly more legal words, had a significantly lower intrusion rate, significantly shorter first-response latencies and final silence periods, as well as significantly shorter between-cluster response latencies. All of the effect sizes but the one for final silence were strong. Additionally, subjects displayed significantly shorter within-cluster compared between-cluster response latencies on both the animal and tree tasks (Z = 119.0, p < .001, r_b_ = 0.983 and Z = 120.0, p < .001, r_b_ > .999, respectively). The number of legal words on both the animal and tree tasks was significantly positively correlated with cluster magnitude (ρ = .547, p = .035 and ρ = .753, p = .001, respectively). No other correlations were significant.

**Table 1.**
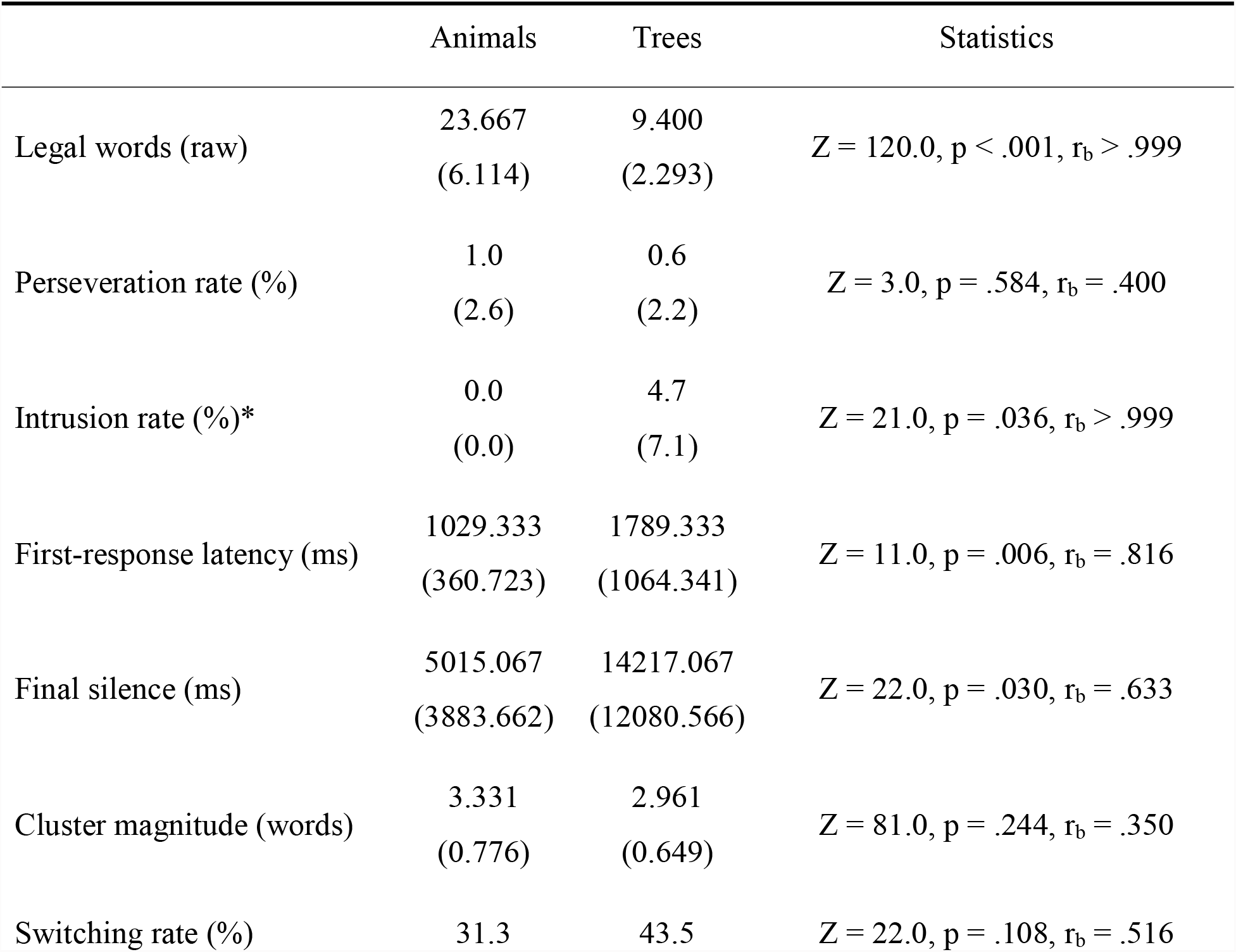
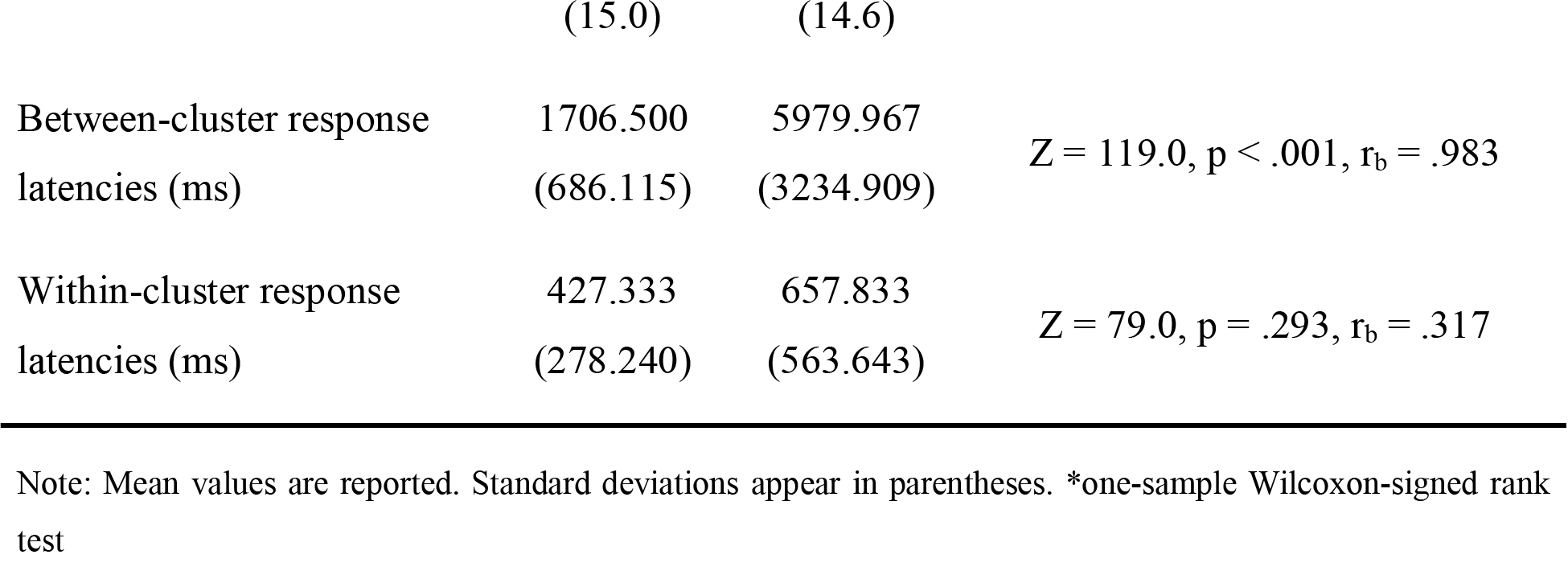
Within-sample comparisons between the animal and tree tasks

### 3.2. Comparisons and correlations within phonemic fluency tasks

Results from the comparisons are displayed in Table 2. On the K compared to the M task, subjects produced significantly more legal words and had significantly shorter between-cluster response latencies. Additionally, there were no significant differences between the between-cluster and within-cluster response latencies on either the K or M task (Z = 31.0, p = .569, r_b_ = .205 and Z = 25.0, p = .301, r_b_ = .359, respectively).

**Table 2.** Within-sample comparisons between the K and M tasks

|  | K | M | Statistics |
| --- | --- | --- | --- |
| Legal words (raw) | 15.750<br>(2.864) | 12.833<br>(4.086) | $Z = 58.0, p = .029, r_b = .758$ |
| Perseveration rate (%) | 0.4<br>(1.3) | 1.2<br>(2.9) | $Z = 0.0, p = .371, r_b > .999$ |
| Intrusion rate (%) | 0.0<br>(0.0) | 0.0<br>(0.0) | N/A |
| First-response latency (ms) | 2409.167<br>(1709.005) | 1893.333<br>(640.289) | $Z = 50.0, p = .424, r_b = .282$ |
| Final silence (ms) | 3740.500<br>(3270.119) | 6285.333<br>(6335.809) | $Z = 30.0, p = .519, r_b = .231$ |
| Cluster magnitude (words) | 2.562<br>(0.770) | 2.540<br>(0.830) | $Z = 25.0, p = .813, r_b = .111$ |
| Switching rate (%) | 72.3<br>(13.9) | 71.4<br>(11.4) | $Z = 29.000, p = .756, r_b = .121$ |
| Between-cluster response latencies (ms) | 2014.583<br>(787.570) | 2732.708<br>(1158.775) | $Z = 9.0, p = .016, r_b = .769$ |
| Within-cluster response latencies (ms) | 1864.375<br>(1347.603) | 2169.375<br>(1698.251) | $Z = 39.0, p > .999, r_b < .001$ |

Note: Mean values are reported. Standard deviations appear in parentheses.

The number of legal words on the K task was significantly positively and strongly correlated with cluster magnitude (ρ = .731, p = .007), while on the M task none of the correlations were significant. No other correlations were significant.

### 3.3. Correlations with sociodemographic variables

As expected in a sample of university students, age and education were significantly positively and strongly correlated (ρ = .746, p < .001).

Both age and education were significantly positively and moderately correlated with cluster magnitude on the animal task (ρ = .659, p = .008 and ρ = .689, p = .005, respectively) but there were no significant correlations between the tree task and sociodemographic variables.

Age was significantly negatively and moderately correlated with the first-response latency on the K task (ρ = -.623, p = .030). There were no significant correlations between the M task and sociodemographic variables.

## 4. DISCUSSION

### 4.1. Semantic fluency

Comparisons between the animal and tree fluency tasks revealed highly different performance patterns across the two tasks. Subjects produced significantly 60.28 % more legal words on the animal compared to the tree category, suggesting that the subjects in our sample had access to more animal than tree words, as well as that the animal task was appreciably less difficult compared to the tree task. Both the former and the latter suggestions are also arguably indicated by the significant differences in the duration of the final silence between the two tasks. On average, subjects stopped responding in the 55. second on the animal task compared to the 46. second on the tree task, with the latter possibly indicating that the readily available tree words were more or less exhausted before the end of the task and that the access to less familiar (but entrenched) words was much more demanding in the tree compared to the animal task. Subjects were further significantly slower to utter the first response on the tree compared to the animal task, indicating that it was more difficult for the subjects to access the category *trees*, retrieve, and utter the first tree word compared to the category *animals*. The results regarding first-response latency and final silence indicate that the tree task was more difficult for the subjects compared to the animal task during both the incipient and ending stages of the task.

Significant differences between the two tasks were identified in the intrusion rate as well. While no subjects produced intrusions on the animal task, intrusions on the tree task were produced by seven subjects. We would like to note at this point that we were very careful in classifying responses as intrusions and that our criteria for intrusion classification were more lax (but nevertheless precise) compared to, for example, Moreno-Martínez et al. (2017) who classified fruit trees (such as *pear*) as intrusions on the tree task. In our study, fruit trees were classified as legal because we found there was no reason to classify them as errors. Intrusions on the tree task included: *glog* ‘hawthorn’ (2), *bugenvilija* ‘bougainvillea’ (1), *hortenzija* ‘hortensia’ (1), *ebanovina* ‘ebony’ (1), *ruža* ‘rose’ (1), *jorgovan* ‘lilac’ (1), *loza* ‘grapevine’, and *žir* ‘acorn’ (1). The significantly higher intrusion rate on the tree compared to the animal task indicates that the boundaries of the category *trees* were considerably looser compared to the boundaries of the category *animals* in our sample.

Subjects did not exhibit significant differences in the clustering and switching strategies between the two SF tasks. However, while descriptive data showed only slight differences in cluster magnitude, the differences in the switching rate were more pronounced, suggesting that subjects were less likely to produce a cluster once a switch has been uttered on the tree compared to the animal task. This might reflect lesser functional connectivity between concepts in the category *trees* compared to *animals* and, specifically, lesser hypernymic/hyponymic organization within the category *trees* compared to *animals*. In any case, the existence of clusters on the tree task still indicates the presence of hypernymic/hyponymic and co-hyponymic structures in the category *trees*. Subjects were significantly slower in between-cluster response latencies on the tree compared to the animal task indicating that the functional connectivity between subcategories is less efficient in the category *trees* compared to *animals*. Interestingly, larger clusters were associated with better productivity on both the animal and tree tasks, yet the correlation coefficient was considerably higher in the tree task. Thus, those who had better organization of hypernymic/hyponymic and co-hyponymic relations between concepts in both categories produced more words on the given SF task and this effect was more pronounced on the animal task.

### 4.2. Phonemic fluency

The differences in performance patterns between the two PF tasks were much less pronounced than between the two SF tasks. Significant differences were observed in the productivity and between-cluster response latencies, with subjects producing significantly more words and having significantly shorter between-cluster response latencies on the K compared to the M task. These results suggest that performance on the K task was easier for the subjects compared to the M task. Because there are presumably more words starting with ⟨m⟩ compared to the ⟨k⟩ task, this result presumably does not reflect the disproportionate number of words starting with either ⟨k⟩ or ⟨m⟩ in the Croatian dialects.

## 5. CONCLUSION

Although preliminary, our results potentially have important implications for neuropsychological research utilizing VF tasks. VF studies typically draw the same inferences from different categories or letters and in the case of using multiple categories or letters, the aggregate result of SF and LF is typically calculated and used in analyses. Our results show that performance may vary depending on the given category or letter and we argue that this may reflect differential linguistic processes. We further find it likely that these disproportionalities also reflect differential cognitive processes, as those assessed by standard neuropsychological tasks used in psychiatric and neurological research. Thus, we do not agree that the same inferences can be drawn from different categories or letters. Given the frequent use of VF in psychiatry, neurology, and other disciplines, there is an urgent need to reexamine our understanding of VF and its associations with cognitive and neural processes, e.g., by investigating behavioral, cognitive, and neural correlates of different categories and letters and subsequently to revisit previous research.

## Data Availability

All data produced in the present study are available upon reasonable request to the authors.

## Acknowledgments

The content of the manuscript has previously appeared online as a preprint (Gabrić & Vandek, 2021b, 2022). Parts of this research were presented on October 23rd 2020 at the 12th Annual Meeting of the Society for the Neurobiology of Language (Gabrić & Vandek, 2020).

## Ethics statement

Ethical approval was obtained by the Ethical Committee of the Faculty of Humanities and Social Sciences, University of Zagreb (dated 2018/10/03). All subjects signed an informed consent form.

